# COVID-19 Deaths: Which Explanatory Variables Matter the Most?

**DOI:** 10.1101/2020.06.11.20129007

**Authors:** Pete Riley, Allison Riley, James Turtle, Michal Ben-Nun

**Affiliations:** Predictive Science Inc., San Diego, California, USA

**Keywords:** COVID-19, Severe Acute Respiratory Syndrome Coronavirus 2 (SARS-CoV-2), Mechanistic Modeling, SEIRX

## Abstract

As Severe Acute Respiratory Syndrome Coronavirus 2 (SARS-CoV-2) spreads around the World, many questions about the disease are being answered; however, many more remain poorly understood. Although the situation is rapidly evolving, with datasets being continually corrected or updated, it is crucial to understand what factors may be driving transmission through different populations. While studies are beginning to highlight specific parameters that may be playing a role, few have attempted to thoroughly estimate the relative importance of these disparate variables that likely include: climate, population demographics, and imposed state interventions. In this report, we compiled a database of more than 28 potentially explanatory variables for each of the 50 U.S. states through early May 2020. Using a combination of traditional statistical and modern machine learning approaches, we identified those variables that were the most statistically significant, and, those that were the most important. These variables were chosen to be fiduciaries of a range of possible drivers for COVID-19 deaths in the USA. We found that population-weighted density (PWD), some “stay at home” metrics, monthly temperature and precipitation, race/ethnicity, and chronic low-respiratory death rate, were all statistically significant. Of these, PWD and mobility metrics dominated. This suggests that the biggest impact on COVID-19 deaths was, at least initially, a function of where you lived, and not what you did. However, clearly, increasing social distancing has the net effect of (at least temporarily) reducing the effective PWD. Our results strongly support the idea that the loosening of “lock-down” orders should be tailored to the local PWD. In contrast to these variables, while still statistically significant, race/ethnicity, health, and climate effects could only account for a few percent of the variability in deaths. Where associations were anticipated but were not found, we discuss how limitations in the parameters chosen may mask a contribution that might otherwise be present.

## 1. Introduction

Following the first signs of its presence in December 2019 (Huang *and others*, 2020), COVID-19 has rapidly spread across the world, impacting almost every country in its wake (Dong *and others*, 2020). However, its impact has been markedly different (at least as of late May) in different locations, a consequence of factors that remain actively debated. We can categorize these factors as follows. First, imposed societal interventions, in the form of social distancing measures (e.g., “shelter in place” orders, self-isolation, school closures, etc.). Second, demographic differences in the population including age distribution, health, gender, and race of the populations. Third, climatic variability (e.g., temperature, precipitation, or humidity variations), and spatial heterogeneity in terms of the density of people that a person is typically exposed to. Fourth, environmental variability such as pollution. Fifth, virus connectivity factors, such as the likelihood that the disease will be imported into an area from outside. These are not mutually exclusive categories, and, indeed, we would anticipate a number of confounding variables amongst them.

Our objective in this study is to assess which of these explanatory (independent) variables are clearly important for predicting COVID-19 deaths, primarily to aid in the development of mechanistic model refinements (Riley *and others*, 2013, 2015, 2017; Ben-Nun *and others*, 2019; Turtle *and others*, 2019). Mechanistic models, as opposed to statistical or machine-learning models are typically developed by incorporating the minimum number of underlying processes necessary to describe the observations sufficiently well to answer specific questions. The types of questions being asked about COVID-19 include, but are not limited to: What impact will interventions have on the trajectory of the disease? Will weather conditions cause the disease to (at least temporarily) abate as we approach the northern summer? Which locations are likely to suffer the most severe outcomes? Which populations will likely fair better, and which will fair worse? A crucial step in answering these questions using models, then, is to understand what factors appear to matter.

In this report we assemble an illustrative dataset of parameters that capture, at least in principle, these factors. We perform several multiple-linear-regression (MLR) and machine learning (ML) analyses on the data to identify the key explanatory variables that are likely driving the variations in the number of deaths per 100,000 for the 50 contiguous U.S. states (i.e., their statistical significance). Additionally, we assess the degree to which these variables can explain the observed deaths (i.e., their importance).

## 2. Methodology

### 2.1 Data

Data were collected from a variety of sources, each chosen as a fiduciary of one of the factors described above. They are summarized in Table 1. Briefly, the first six parameters (retail, grocery, parks, transit, workplaces, and residential) were scraped from Google’s Mobility Reports on April 20 (Google), and represent the fractional change (expressed as a percentage) in people’s travel to these locations. Thus, a mean value of 11.38% in ‘residential’ represents the increase in people remaining at home since the start of “shelter in place” orders. Similarly, a decrease of 46.58% in ‘retail’ reflects the drop in people going to retail locations. Many of the parameters are self-explanatory, including: age, % 65 years and over (USCensus), obesity rates (LiveScience), chronic lower respiratory death rates (CDC), pollution indices (USNews), the date that a state of emergency was declared (Wikipedia), average relative annual and spring-time humidity, dew point, temperature, precipitation (Brettschneider; CurrentResults), and UV index (USEPA). However, several require further explanation.

**Table 1.** List of parameters used in multiple-regression analysis, together with their basic statistical properties. See text for detailed explanation of each parameter.

| Statistic | N | Mean | St. Dev. | Min | Pctl(25) | Pctl(75) | Max |
| --- | --- | --- | --- | --- | --- | --- | --- |
| Retail | 50 | -46.580 | 6.081 | -66 | -50 | -42.2 | -36 |
| Grocery | 50 | -17.380 | 7.982 | -34 | -23.8 | -11 | 1 |
| parks | 50 | 16.400 | 42.524 | -70 | -6 | 29.8 | 134 |
| Transit | 50 | -48.520 | 13.167 | -76 | -58 | -37.5 | -21 |
| workplaces | 50 | -37.760 | 5.336 | -55 | -41.8 | -35 | -27 |
| Residential | 50 | 11.380 | 2.147 | 8 | 10 | 12 | 16 |
| Age | 50 | 38.616 | 2.328 | 31.000 | 37.325 | 39.600 | 44.900 |
| Low indust. toxins | 50 | 25.500 | 14.577 | 1 | 13.2 | 37.8 | 50 |
| Low poll. health risk | 50 | 25.500 | 14.577 | 1 | 13.2 | 37.8 | 50 |
| Chron. low resp. death rate | 50 | 42.790 | 10.520 | 20 | 34.9 | 48.7 | 64 |
| $\geq 65$ years old | 50 | 16.506 | 1.926 | 11 | 15.7 | 17.5 | 21 |
| Race param. 1 | 50 | 11.954 | 9.686 | 0.900 | 4.850 | 15.975 | 38.900 |
| Race param. 2 | 50 | 5.580 | 8.046 | 1.100 | 2.325 | 5.775 | 56.800 |
| Race param. 3 | 50 | 12.070 | 10.469 | 1.400 | 5.125 | 13.975 | 49.100 |
| Race param. 4 | 50 | 76.164 | 12.881 | 24.300 | 67.825 | 85.000 | 94.300 |
| Obesity rates | 50 | 27.740 | 3.305 | 19.000 | 25.600 | 29.600 | 35.200 |
| Av. rel. humidity | 50 | 67.102 | 8.346 | 38.300 | 65.950 | 71.475 | 77.100 |
| Av. dew point | 50 | 41.710 | 9.224 | 26.500 | 34.925 | 46.750 | 65.200 |
| Av. ann. temp. (C) | 50 | 11.074 | 4.830 | -3.000 | 7.325 | 14.800 | 21.500 |
| Av. ann. precip. (mm) | 50 | 941.820 | 371.752 | 241 | 622.5 | 1,216.2 | 1,618 |
| State of emerg. dec. | 50 | -3.100 | 3.666 | -13 | -4.8 | 0 | 3 |
| Av. spring temp. | 50 | 10.550 | 4.941 | -4.100 | 6.650 | 14.050 | 21.100 |
| Av. spring precip. | 50 | 82.840 | 34.067 | 20 | 56.8 | 104.5 | 151 |
| Rel. hum. (morning) | 50 | 76.280 | 8.593 | 43 | 73.2 | 81 | 90 |
| rel. hum. (afternoon) | 50 | 48.440 | 9.004 | 17 | 47 | 53.8 | 61 |
| UV Index | 50 | 7.780 | 2.376 | 1 | 7 | 9 | 12 |
| Deaths/100k ($n_{deaths}$) | 50 | 20.769 | 52.440 | 0.092 | 1.313 | 18.250 | 349.854 |
| PWD | 50 | 3,298.370 | 4,113.740 | 694.900 | 1,356.450 | 3,561.875 | 28,161.500 |
| Date of first death | 50 | 18,341.040 | 6.465 | 18,331 | 18,337 | 18,346 | 18,365 |

Population-weighted density (PWD) is, the average of each resident’s census tract density, effectively, the density at which the average person lives. It is important to distinguish this from the average population density for a location if there is any significant heterogeneity in population density. For example, the population density of the entire U.S. is approximately 90 people per square mile. In contrast, the population-weighted density is more than 5,000 people per square mile. These data were derived from the WorldPop dataset (Worldpop).

The reported date of the first death in each state is used as a proxy for initial seeding of the virus in each state, and, by extension, a proxy for the connectivity of the state to likely sources of the disease (e.g., China, Europe, or other states).

Race parameters 1 through 4 are an attempt to identify any possible links with the underlying races/ethnicities within each state (USCensus). Specifically, they are: (1) Percent Estimate-Race alone or in combination with one or more other races-Black or African American; (2) Percent Estimate-Race alone or in combination with one or more other races-Asian; (3) Percent Estimate-Hispanic or Latino (of any race); and (4) Percent Estimate-One race-White (Smith, 2014).

Finally, our response (dependent) variable is the number of deaths per 100,000 (JHU). All values reported here are correct through May 10 2020. This is, of course, an ever-changing value; however, both the cumulative number of confirmed cases as well as the number of deaths per 100,000 for each state tended to maintain similar relative slopes (Figure 1), such that the results do not depend sensitively on the precise date chosen to represent the number of deaths in each state. In fact, the analysis was repeated for five different dates from April 25 through May 10 with no qualitative difference in the results, or their interpretation. It is, nevertheless, worth noting several outliers. Washington, in particular, was the first state to report an appreciable number of deaths in mid-March; however, its initial slope remained shallower than other states, and, by early April, became even flatter. The net effect of this is that in terms of relative number of deaths, from early April to early May, it exchanged position with a dozen other states. The relative positions of other states has also changed during the course of the last two months. In general, however, the changes were likely driven by a particular event that changed the evolving slope of the profile at some point. These typically occurred in the early portion of the outbreak, and the relative positions of the states in the last month has remained largely unaltered.

**Fig. 1.**
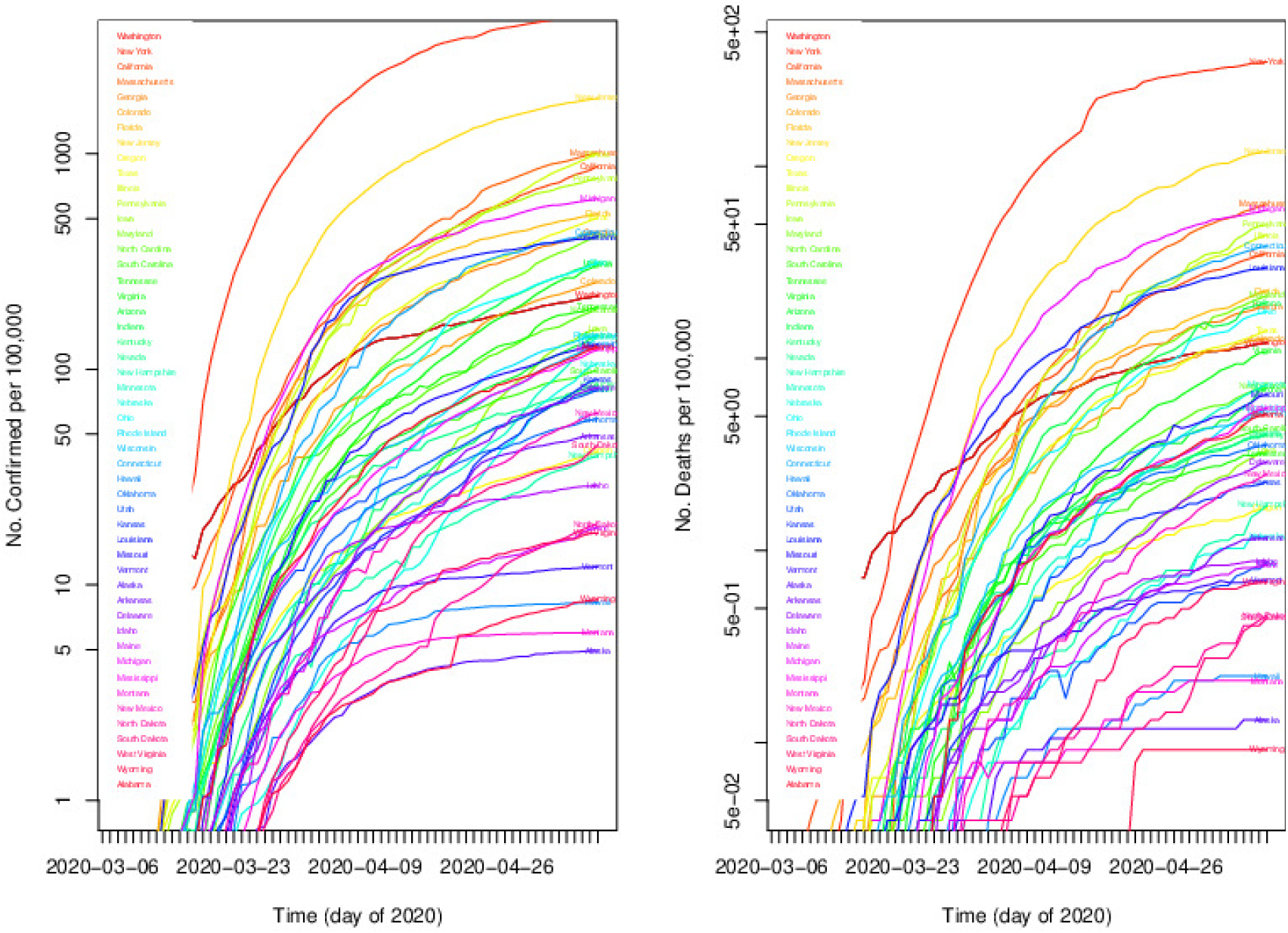
The variation of (Left) Cumulative number of confirmed cases per 100,000 for each state (colored arbitrarily to better separate each curve). (Right) Cumulative number of deaths per 100,000 for each state. Data runs from 2020-03-06 through 2020-05-10.

### 2.2 Models

Multiple regression models were analyzed using several packages in *R*, primarily relying on ‘lm’ (R Core Team, 2019). To identify the explanatory variables that provided the best performing model (i.e., one that lowered the prediction error), we used the ‘stepAIC’ function from the MASS package (Venables and Ripley, 2002). Both forward selection and backward selection (i.e., backward elimination) were performed (James *and others*, 2013). In the former, initially, no predictors are included in the model, and the algorithm iteratively adds the most contributory variables, stopping when the improvement is no longer statistically significant.

These regression techniques, however, only identify the statistical significance of the variables. To estimate the relative importance of each explanatory variable in describing the variability in deaths, we applied a number of traditional statistical and more modern machine learning (ML) approaches. Using multiple approaches is important for assessing the uncertainty that should be ascribed to a particular ordering of the variables, since different techniques rely on different metrics for importance. The techniques applied included: random forest (Liaw *and others*, 2002), Xgboost (Chen and Guestrin, 2016), relative importance (Grömping *and others*, 2006), earth (wrapper, 2019), step-wise regression (Bendel and Afifi, 1976), and DALEX (Biecek, 2018). It is important to underscore that these techniques use different definitions of what signifies”important”, and, thus, we do not expect agreement between the results. Nevertheless, where the results do agree is where we can be most confident that the explanatory variable importance is significant, and where they do not, we must remain more cautious.

## 3. Results

When a multiple linear regression analysis was performed on the 28-parameter dataset, four explanatory variables were found to be significant: retail, average annual precipitation, average spring precipitation, and PWD (Table S1). Together, *R*^2^ for these variables was 0.973, and thus, these variables are capable of explaining 97% of the variations in deaths/100,000 (*n*_*deaths*_). We then applied a heuristic, recursive process of elimination to each of the parameters to identify those that, under a more focused analysis might raise to the point of significance. From this process, we identified five explanatory variables: retail, grocery, PWD, average spring temperature and precipitation. Thus, annual precipitation was no longer deemed to be significant, but grocery and average spring temperature now were (Table S2).

Comparison of the five most significant variables with one another showed several illuminating features (Figure S1). First, the number of deaths per 100,000 were clustered below 100 per 100,000, with New York being a significant outlier. Second, unsurprisingly, retail and grocery mobility metrics appear to be the most highly correlated of the explanatory variables. Third, the best fit curve to the number of deaths per 100,000 and PWD appears to (1) be linear, and (2) display the strongest correlation with *n*_*deaths*_.

An analysis of the residuals suggests that Hawaii, New Mexico, and Rhode Island (observations 11, 31, and 39) deviate most (Figure S2 (left)). The fitted line through the residuals suggest the possibility that a non-linear relationship might be more appropriate, however, given the relatively high *R*^2^ *∼* 0.9, we suggest that this is not necessarily so. Comparing the standardized residuals against theoretical quantiles suggests that the errors are approximately normally distributed, at least for most of the states. Notably, New York (32) and Hawaii (11) (and, to a lesser extent, Rhode Island (39)) are outliers (Figure S2 (right)). To check for homoscedasticity, that is, the assumption of equal variance, we can assess whether the residuals are spread equally along the ranges of the predictors. The notable outliers are, once again, Hawaii (11), Rhode Island (39), and New York (32). Although there is variability, with the exception of New York, the standardized residuals are roughly equal to one and constant along the fitted values (Figure S3 (left)).

Finally, considering the variation of the residuals against leverages, we can estimate to what extent the outliers are influential in the regression analysis. The leverage of a particular observation (state) is related to how much its value on the predictor variable differs from the mean of the predictor variable. We use Cook’s distance as a measure of the influence of an observation, with values lying in the upper-right or lower-right being indicative of unduly influential observations. Unsurprisingly, New York (32) is clearly influential (Figure S3 (right)). Following this, Hawaii (11), and to an even lesser extent, New Jersey are leveraging the results.

In summary, based on this exploratory analysis, the strongest explanatory variable for deaths per 100,000 is PWD (Figure 2). There is a clear trend for states with increasing PWD to have higher number of deaths. There is, however, substantial scatter away from the smooth line through the data, at least in part due to the contribution(s) from some of the other explanatory variables (as well as potentially other equally, or more important variables not included). For example, it is worth noting that, geographically, the states substantially below the line are located at higher latitudes and away from either coast (N. and S. Dakota, Wyoming, Utah, Nebraska, Nevada). On the other hand, many of the states substantially above the line are from southern regions (Mississippi, Alabama, S. Carolina, Kentucky, Missouri, Louisiana). It could also be argued that this demarcation is reflective of other factors, including political affiliation, which may have modulated the timing and/or efficacy of the “shelter in place” orders. Finally, we remark that Hawaii, and to a lesser extent (because of the log-log scale) Alaska, are unique outliers in terms of unusually low deaths given their relative PWD.

**Fig. 2.**
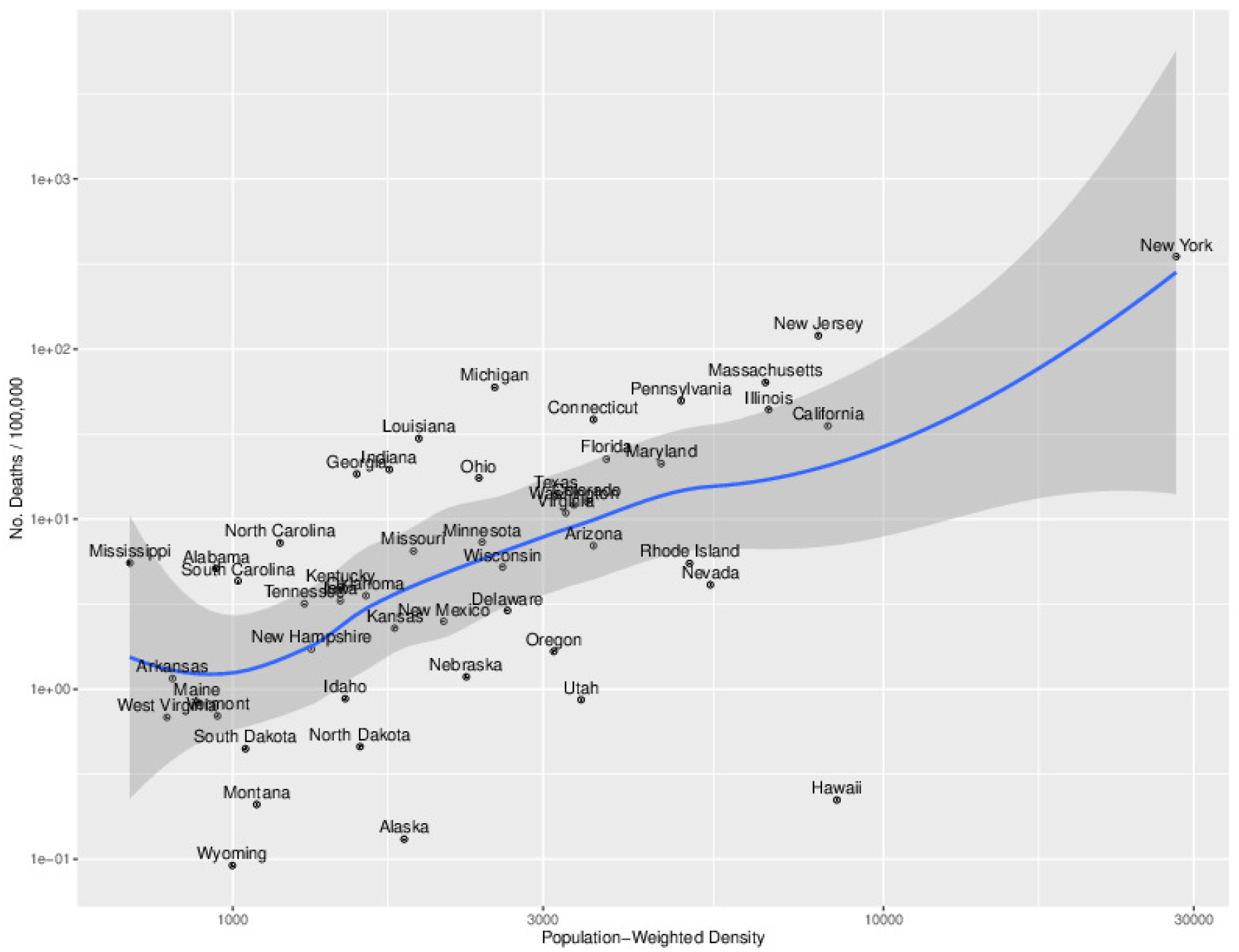
The relationship between the number of deaths per 100,000 and PWD. Each state is identified with a dot and the variability and smooth profile are shown with the dark grey region and blue line, respectively.

While the preceding analysis heuristically investigated the possible contribution of the explanatory variables in describing the variations in *n*_*deaths*_, we next apply more robust techniques using a variety of algorithms to assess both statistical significance and importance. In these approaches, explanatory variables are iteratively added or removed to identify the subset of variables that produce the best performing model, that is, the model with the lowest prediction error.

First, using *R*’s MASS package, we applied both forward selection, where variables were added iteratively, until the improvement is no longer statistically significant, and backward elimination, where variables are iteratively removed until the point is reached where all the variables are statistically significant (Table S3). Based on this analysis, 14 variables can account for 97% of the variability in *n*_*deaths*_. This is approximately the same as including all 28 variables, and substantially higher than the 87.5% that we computed using our *ad hoc* approach of searching through the variables. PWD, race parameter 1, average annual temperature and precipitation, average spring precipitation are the strongest contributors.

Using the Random Forest method to assess the importance of the variables, we found (in order of importance): PWD, retail, chronic low-respiratory death rate, residential, race parameter 3, obesity rates, average spring temperature, average dew point, race parameter 1, and date of first death were the most important (Table S4). Comparing this list with the significant variables identified earlier, suggests that any subset of significant *and* important explanatory variables would include at least PWD, retail, chronic low-respiratory death rate, race parameter 1, and date of first death.

Applying the Xgboost method for ranking explanatory variables in order of their importance identified the ‘retail’ mobility metric as the most important variable, followed by PWD, and then two more mobility metrics (workplaces and grocery). Race parameter 1, relative humidity (afternoon), date of first death, and average spring temperature and dew point followed. Finally, Chronic low-respiratory death rate was the remaining variable but only 1% as important as the most important variable (Table S5).

The Multivariate Adaptive Regression Splines (MARS) model can also be used to rank explanatory variables. Unlike the random forest, it has been shown to be more susceptible to unstable explanatory variables. Nevertheless, it is a flexible technique and is included here to provide evidence for the sensitivity of our results to the technique implemented. PWD, race parameter 2, and retail captured more than 90% of the importance (Table S6), with grocery and averge annual precipitation rounding out the top-five variables.

The step-wise regression method can also be combined with the Akaike Information criteria (AIC) to identify the best model, that is, the best combination of parameters to explain the output variable. Using this approach, in order of importance, PWD, grocery, retail, average spring temperature and precipitation were all found to be important (Table S7).

Finally, we consider the DALEX package, which is, in fact, a meta-package in the sense that it can compare responses from different models to allow for direct comparison. Here, though, we apply the DALEX machinery with the random forest technique and use the ‘explain’ and ‘variable importance’ functions to quantify their relative importance (Table S8). PWD, retail/workplaces, chronic low respiratory death rate, average relative humidity, grocery, and race parameter 1 describe most of the observed variability in *n*_*deaths*_. The degree to which these variables contribute to the overall variations can also be visualized graphically (Figure 3). Thus, we infer that the first four variables account for most of the observations. Of these, however, average relative humidity was not found to be statistically significant in any of our analyses. Thus, while humidity was “important”, it wasn’t “significant”.

**Fig. 3.**
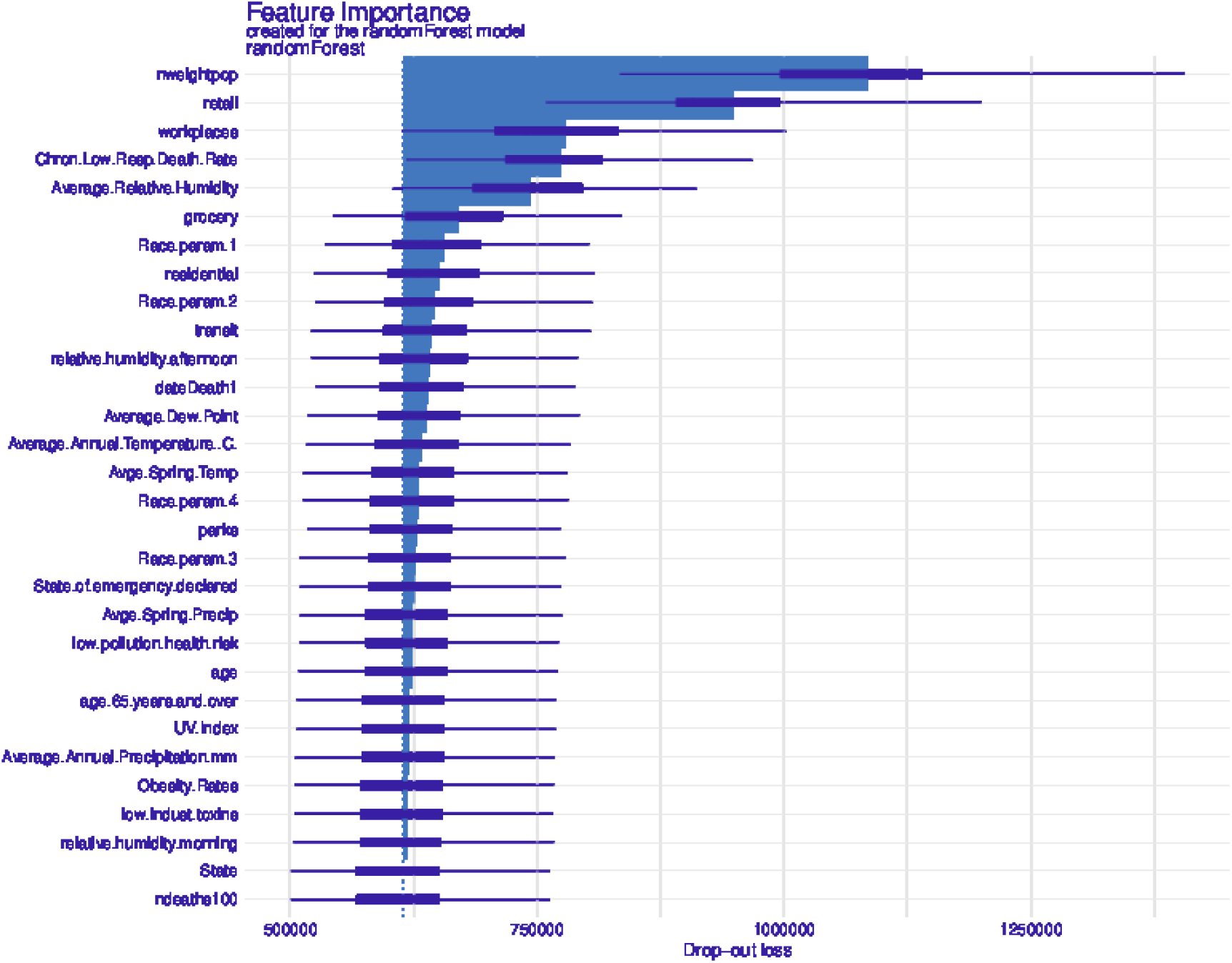
Relative importance of explanatory variables using the DALEX method. See text for more details.

## 4. Discussion

In this study, we have presented an illustrative, but not exhaustive multiple-regression analysis of 28 explanatory variables, in an attempt to identify those parameters that can best predict deaths per capita in U.S. states. Our results are not surprising in the sense that we would have anticipated that PWD and lockdowns, in particular, would likely explain at least some of the variations in deaths. It is also reassuring that our results support other studies highlighting the association of race/ethnicity (Yancy, 2020), climatic conditions (Sajadi *and others*, 2020), and population demographics (Flaxman *and others*, 2020*a*) with deaths. In fact, given the relative homogeneity of the U.S. population, it is somewhat remarkable that small variations from state to state would result in measurable and statistically-significant results.

Our initial dataset contained a number of other variables that were dropped from the formal analysis, primarily because, although there might be an obvious association with deaths, there was no conceivable driving mechanism (i.e., causal relationship). For example, we found that the political affiliation of each state (as measured by the results of the 2016 election) was a strong and statistically significant explanatory variable. In addition to this simply being a confounding variable for several others, such as PWD (republican states tend to be more rural) and the efficacy of lockdowns (democratic states typically implemented orders earlier and/or more stringently), its variation serves no useful role in explaining the number of deaths. Nevertheless, we emphasize the point here to highlight the dangers in over-interpreting what, based on the analysis presented here, can only be considered associations.

Making the leap from associations to causal relationships can be accomplished by mathematical models. Thus, we suggest that the results presented here can be further tested within various types of mechanistic models. Indeed, the two major associations of PWD and mobility metrics are, to varying degrees, already incorporated into models. Individual-based spatial models, for example, can explicitly account for the population-weighted density of the population as well as (at least parametrically) the mobility of people (Flaxman *and others*, 2020*b*). The significance of climate (particularly temperature, precipitation, and/or humidity) can readily be incorporated as well (Riley *and others*, 2017; Ben-Nun *and others*, 2019; Turtle *and others*, 2019).

It is not surprising that PWD was the most important explanatory variable. It also must be recognized that this variable is related to each state’s social distancing orders, with the primary difference that PWD is generally immutable, whereas social distancing can and will change over time. It may be tempting, particularly for vulnerable individuals to move to a region of lower PWD, as this will likely have a strong impact on their likelihood of dying from COVID-19. It could be argued that this should be balanced with the likelihood of the quality of healthcare being proportionately worse in more rural areas. However, based on the simple, but clear relationship between the number of deaths per 100,000 and PWD, this appears not to be the case: Regions of lower PWD are experiencing lower deaths per capita. Moreover, based on the roughly constant gradients (Figure 1(b)), this has been true for the past two months, and will likely hold for the foreseeable future.

Although it is intuitively obvious that PWD and *n*_*deaths*_ should be positively correlated in terms of causality, increases in PWD would primarily drive higher incidence, which would, in turn, result in more deaths. However, an interesting question not addressed here is whether the relationship between PWD and deaths is linear? If a disproportionate number of more vulnerable people live in higher-PWD regions, this would result in a plot of *n*_*deaths*_ versus PWD curing upwards. There is some suggestion of this in our results (Figure 2); however, such an inference is at best tentative and requires more detailed analysis, including additional datasets that are likely not yet available.

It was somewhat surprising that no significance was found for any measures of age, chronic disease, pollution, or some of the race parameters. In some cases, this is likely due to the fact that the variability was just too small to drive a signal in the results. For example, while the average obesity rate across the 50 states is 27.7%, the standard deviation is only 3.3%. In these cases, we suggest that rather than there not being contribution, we were unable to identify it because of the quality of the parameter. Indeed, several recent studies, which have focused specifically on a particular parameter, such as race or ethnicity, were able to identify it within smaller populations with higher heterogeneity (Raifman and Raifman, 2020). Additionally, where explanatory variables are co-linear, the explanatory capability might be absorbed by another parameter.

Equally remarkable is that small differences in race/ethnicity amongst states were statistically significant, albeit only accounting for a small fraction in the variability of the deaths. This suggests that these factors may be more important than those that were not found to be statistically significant here. Of course, this remains speculation until more definitive studies are completed.

## 5. Conclusions

In this study, we have applied MLR and ML analyses to a large number of potentially promising explanatory variables. We found that population-weighted density (PWD), as well as metrics for the efficacy of lock-down orders were the most significant explanatory variables in explaining the variability in the rate of deaths across U.S. states. Race/ethnicity, health, and climate effects, while statistically significant, were not as significantly important. This may reflect an intrinsic truth, or, more, more likely, is a consequence of the limitations in the data collected. Further studies with more extensive and accurate datasets (as they become available) will likely resolve these issues.

## 6. Software

Software in the form of R code, together with a sample input data set and complete documentation is available on request from the corresponding author.

## Acknowledgments

This work was supported through a RAPID grant from the National Science Foundation (Award Number (FAIN): 2031536).

